# The experience of distress during the COVID-19 outbreak: a cross-country examination on the fear of COVID-19 and the sense of loneliness

**DOI:** 10.1101/2020.11.24.20237586

**Authors:** Gianluca Lo Coco, Ambra Gentile, Ksenija Bosnar, Ivana Milovanovic, Antonino Bianco, Patrik Drid, Saša Pišot

## Abstract

**Objectives:** To examine gender, age and cross-country differences in fear of COVID-19 and sense of loneliness during the lockdown, by comparing people from countries with a high rate of infections and deaths (i.e. Spain and Italy) and from countries with a mild spread of infection (i.e. Croatia, Serbia, Slovakia, Slovenia, Bosnia and Herzegovina).

**Methods:** A total of 3876 participants (63% female) completed an online survey on “Everyday life practices in COVID-19 time” in April 2020, including measures of fear of COVID-19 and loneliness.

**Results:** Males and females of all age groups in countries suffering from a strong impact of the COVID-19 pandemic reported higher fear of COVID-19 and sense of loneliness. In less endangered countries females and elder stated more symptoms than males and younger; in Spanish and Italian sample the pattern of differences is considerably more complex.

**Conclusion:** Future research should thoroughly examine different age and gender groups. The analysis of emotional well-being in groups at risk of mental health issues can help to lessen the long term social and economic costs due to the COVID-19 outbreak.

## Introduction

By the mid of January, the Chinese government quarantined the city of Wuhan (11 million inhabitants) and subsequently extended the measure to Hubei province (60 million inhabitants) to contain the Coronavirus Infectious Disease 2019 (COVID-19) epidemic. Since that time, there has been a progressive spread of the virus throughout the world, infecting millions of people and causing hundreds of thousands of deaths. On March 11, 2020, the World Health Organization (WHO) declared a state of pandemic. Quarantine (i.e. the segregation of one or more healthy people inside their own homes, to prevent infection and the virus spreading) was considered one of the most helpful measures in containing the infection. Most countries issued varying degrees of “shelter-in-place” orders [1] and almost one-third of the global population faced some form of quarantine due to the COVID-19 outbreak in the last few months. However, there is evidence that facing a quarantine can have detrimental effects on people’s psychological health [2], with anxiety, anger, insomnia, somatic symptoms, mainly due to the loss of freedom, the separation from loved ones, uncertainty over the disease, shortage of living supplies.

To date, there is some initial evidence showing the negative impact of the COVID-19 pandemic on psychological well-being [3-5]. One of the first surveys, which was conducted in China during the lockdown showed that more than 50% of participants rated the psychological impact of COVID-19-related restrictions as moderate or severe [6], with greater difficulties associated with the effects of COVID-19 pandemic on daily life and social and work activities [7-9]. Of course, this negative impact is even stronger for healthcare professionals facing this global crisis [10], with a considerable proportion of workers reporting symptoms of depression, anxiety and stress [11]. The negative psychological effects of the COVID-19 pandemic on the individual’s mental health states were further confirmed in studies from several Western countries [12-14].

As COVID-19 continues to spread, so does the research on people’s experience of fear during the pandemic. Fear of personal infection or infection of loved ones is common among people exposed to any infectious disease outbreak [2, 9], and a specific examination of the characteristics of the fear of COVID-19 is worthwhile. Globally, more than 26 million people contracted the virus infection, and 870,000 have died. Thus, it is likely that the high mortality rates due to COVID-19 have negatively impacted on the individual’s feelings of fear of contagion and anxiety across countries all over the world. In the current study, we will focus on a cross-country examination of the COVID-19 outbreak on the fear of COVID-19, by differentiating European countries which reported a severe impact of the infection (i.e. Italy and Spain) and those which showed a mild impact of the infection among populations (i.e. Croatia, Serbia, Slovakia, Slovenia, Bosnia and Herzegovina).

Although fear is an adaptive response in the presence of danger, it was suggested that the construct of fear of COVID-19 should be examined within an integrated complex model [15]. For example, fear of infection can trigger healthy behaviors or, on the contrary, foster health anxiety. Fear of COVID-19 can improve safety behaviors towards the elderly, but can also result in inappropriate worries about loved ones. Concerns and fears about one’s own health and the well-being of one’s own beloved ones (particularly elderly or people suffering from any physical illness) can exacerbate feelings of anxiety. If these concerns are prolonged in time, they may increase the risk of serious mental health conditions, including anxiety disorders, stress and trauma-related disorders [16].

Moreover, feelings of uncertainty about the future and the lack of an effective vaccine may have led people to heighten their fear of COVID-19 during the quarantine. To date, some new tools for the assessment of the Fear of COVID-19 have been developed [17-19] to provide healthcare professionals with a valid measure for monitoring fear and anxiety of individuals during the COVID-19 crisis [17, 20]. Previous research showed a significant association between the fear of COVID-19 and the most recommended strategies to control the spread of COVID-19, such as spatial distancing and handwashing [21, 22]. People with an excessive fear of the infectious outbreak are more likely to report greater psychosocial distress, whereas people showing under-response beliefs and little anxiety are more likely to disregard the physical distancing [19]. There is also evidence that intolerance of uncertainty, risk for loved ones, health anxiety was associated with the fear of COVID-19 [23].

An important step towards understanding the critical characteristics of this construct is to examine the cross-country similarities and differences in fear of COVID-19. Although there is some evidence to suggest that fear of COVID-19 can be concentrated in regions with the highest reported COVID-19 cases [24], limited research efforts examined whether fear may differ in European populations facing a high or limited impact of the infection and severity of restriction policies. Moreover, the association between fear of COVID-19 and social isolation during the lockdown needs to be further investigated in cross-cultural research. To date, the link between people’s experience of fear of COVID-19 and feelings of loneliness received little research attention. Although physical distancing measures have been critical to contain the rate of infection, there is concern that limits on social activities and restrictions on in-person social contacts may increase feelings of loneliness [25, 26]. Prior research on the experience of loneliness in response to the social restrictions due to the COVID-19-related quarantine reported mixed findings. For example, it was shown that being under a stay-at-home order was associated with greater loneliness and health anxiety. However, the higher perceived impact of COVID-19 on participants’ daily life was significantly associated with higher perceived social support and lower loneliness [13]. Moreover, a recent longitudinal study [26] showed no significant changes in loneliness across the three assessment points, between February and the end of April, in USA. Interestingly, despite people perceived an increased absence of social connections during the initial stage of the COVID-19 outbreak, they did not feel more isolated in response of the implementation of social distancing measures. To our knowledge, no previous research examined the link between fear of COVID-19 and feelings of loneliness during the lockdown across countries. Although some studies showed that individuals who felt lonely in the pandemic reported symptoms of anxiety and depression [12, 27], and that greater emotion regulation difficulties and depression may be risk factors for loneliness [28], the interplay between feelings of loneliness and fear of COVID-19 across countries facing varying levels of the spread of infection as well as different home-confinement policies, remains unknown.

The present study examines individual’s experience of fear of COVID-19 and loneliness in response to physical distancing and restriction measures undertaken to contain the outbreak of the COVID-19 in different countries. More specifically, this study aims at examining potential cross-country differences in measures of fear of COVID-19 and loneliness across two groups of European countries with varying impact of the COVID-19 pandemic (e.g., with regards to the number of deaths and measures of total lockdown). Specifically, we expect that countries reporting a high death and infection rate would display a higher fear of COVID-19, with a decreasing feeling of loneliness, compared to countries reporting a low infection and death rate in the midst of the pandemic. We also aim at examining gender and age group differences across countries. We expect gender differences in fear of COVID-19 and loneliness, and it was hypothesized that females would report more fear of COVID-19 and would feel lonelier than males. Finally, we expect that older adults would feel lonelier and would also display higher fear of COVID-19 compared to younger people across countries.

## Materials and Methods

### Participants and data collection

The sample consisted of 3876 participants (1422 males, 2442 females) from 7 European countries (Italy, Spain, Bosnia and Herzegovina, Croatia, Serbia, Slovakia, Slovenia), whose age was comprised between 18 and 82 years (M= 31.94; SD=12.02). The majority of participants described themselves as female (N= 2442, *M*_age_ = 31.88 years, *SD* = 12.96), 1422 described themselves as male (*M*_age_ = 32.05 years, *SD* = 13.14), and 12 described themselves as other gender (e.g., transgender, bigender, non-binary). However, given the low number in this grouping, we limited the analyses to men and women. Participants were recruited by the “snowball” approach (personal transmission of the questionnaire via individual e-addresses databases and posting the link to the questionnaire on social networks, official pages of the partners’ organizations, local on-line newspapers). Participants completed an on-line survey “Everyday life practice in COVID-19 time” during the restriction time for COVID-19 pandemic, from April 15, 2020 to April 28, 2020 [29]. Participants had to be 18 years or older and living in the included European countries. They were categorized into four age groups: emerging adults (between 18 and 25 years old), young adults (between 26 and 39 years old), middle-aged adults (between 40 and 60 years old), and older adults (60 years or older) (see Table 1).

**Table 1.** Discriminant Analysis results

| Roots removed | Eigenvalue | Canonical correlation | Wilks' Lambda | Chi-Squared | df | *p* |
| --- | --- | --- | --- | --- | --- | --- |
| 0 | 0.16 | 0.37 | 0.82 | 776.60 | 60 | 0.00*** |
| 1 | 0.04 | 0.18 | 0.95 | 203.01 | 42 | 0.00*** |
| 2 | 0.01 | 0.11 | 0.98 | 69.44 | 26 | 0.00*** |
| 3 | 0.01 | 0.08 | 0.99 | 24.94 | 12 | 0.02* |

(\*\*p<.001; \*p<.01; \*p<.05).

All materials and procedures were reviewed and approved by the consortium of six partners from Science and Research Centre Koper, Slovenia; Faculties of Sport at University of Novi Sad, Serbia; University of Palermo, Italy; University of Zagreb, Croatia; University of Presov, Slovakia, and University of Cadiz, Spain. The study was conducted in accordance with the ethical standards of the Declaration of Helsinki, and all participants signed statements of informed consent to participate in this study. The Ethics Committee of University of BLINDED FOR REVIEW approved this study. Participants were informed that all data would have been processed and managed by the legislation on the protection of personal data and the General Data Protection Regulation (GDPR). They were able to leave the questionnaire at any stage before the submission process. Only surveys with completed mandatory questions were taken into further analysis.

### Measures

The survey was made up of socio-demographic questions (detecting age, gender, education, and nationality), the Fear of COVID-19 Scale [17], and the Three-Item Loneliness Scale [30]. The questionnaires were translated and back-translated to ensure that the wording was appropriate for Spain, Bosnia and Herzegovina, Croatia, Serbia, Slovakia, and Slovenia. The study was conducted in line with the recent recommendations of Swami and Barron [31].

### Fear of COVID-19 Scale

Participants completed the Fear of COVID-19 Scale by Ahorsu et al. [17]. It consists of 7 items with answers on 5-point scale, from completely disagree to agree. It was constructed considering existing scales on fears, expert evaluations and interviews, and it shows very good psychometric properties. Specifically, it has shown stable psychometric properties across countries, with a good reliability (Cronbach alphas: Italy, 0.86; Spain, 0.87; Bosnia and Herzegovina, 0.89; Croatia, 0.85; Serbia, 0.85; Slovakia, 0.83; Slovenia, 0.85).

### Three-Item Loneliness Scale

Loneliness was measured by the 3-item Loneliness Scale by Hughes et al. [30]. It consists of three items detecting the lack of companionship, the feeling of being left out, and the feeling of being isolated from the others, measured on the frequency Hardly Ever, Some of the Time and Often. For the purposes of the present study, the items were treated as three different indicators of feelings of loneliness.

### Data analysis

Descriptive statistics of the total result for The Fear of Covid-19 Scale and items from Three-Item Loneliness Scale were calculated on the total sample and subgroups regarding gender, age and country. Correlational analysis through Pearson’s r was performed to see if loneliness items and fear of COVID-19 were interrelated. Countries were divided in two groups, specifically, the most endangered Italy and Spain (C2) in one group and Slovenia, Croatia, Serbia, Slovakia, and Bosnia and Herzegovina in the other group (C1).

The canonical discriminant analysis of groups defined by age, gender, and country was performed on the total result of The Fear of COVID-19 Scale and items from the Three-Item Loneliness Scale using the Discriminant Function Analysis procedure described by Jennrich [32] in STATISTICA (version 13.0, TIBCO, USA). The significance of the first and subsequent discriminant functions was tested by Wilks’ lambda values at the level of statistical significance p<0,01. Standardized discriminant coefficients and correlations of independent and discriminant variables were determined. The means for the discriminant functions by group (namely, group centroids) were computed; centroids were represented in three two-dimensional Cartesian space.

## Results

### Preliminary results

Descriptive statistics are summarized in Supplement Table 1. Twelve participants who identified as in the “other” gender category were excluded. Therefore, the total sample size was N=3864 for further testing the Fear of COVID-19 and the Loneliness scale. Fear of COVID-19 and loneliness items (lack of companionship, felling left out and feeling isolated) were significantly correlated at p<.01. Independent variables distributions are significantly different from normal because of skewness, but it should not invalidate the discriminant analysis [33, 34].

### Discriminant analysis

Canonical discriminant analysis of groups defined by age, gender and country for the Fear of COVID-19 and the Three-Item Loneliness Scale resulted in three significant discriminants functions (see Table 1), whose discriminant coefficients are represented in Table 2. The first discriminant function is predominantly defined by the result of Fear of COVID-19 (standardized discriminant coefficient= 0.963; correlation with discriminant function= -0.896); feeling less the lack of companionship contributes to a lesser extent (standardized discriminant coefficient= -0.351; correlation with discriminant function= -0.325). The second discriminant function is mainly defined by feeling isolated from the others (standardized discriminant coefficient= -0.709; correlation with discriminant function= -0.926) and tendency to feel more the lack of companionship (standardized discriminant coefficient= -0.313; correlation with discriminant function= -0.689). The third discriminant function is determined by feeling left out (standardized discriminant coefficient= 1.233, correlation with discriminant function= 0.74); partial contributions of other two measures of loneliness are also visible, but to a much lesser extent (standardized discriminant coefficients of feeling the lack of companionship and feeling isolated are -0.489 and -0.430, respectively).

**Table 2.** Standardized discriminant coefficients and correlations with discriminant functions

| Items | Root 1 |  | Root 2 |  | Root 3 |  |
| --- | --- | --- | --- | --- | --- | --- |
|  | S | F | S | F | S | F |
| The Fear of COVID-19 Scale | 0.963 | 0.896 | -0.239 | -0.405 | -0.174 | -0.048 |
| How often do you feel that you lack companionship? | -0.351 | -0.325 | -0.313 | -0.689 | -0.489 | -0.214 |
| How often do you feel left out? | -0.028 | -0.056 | -0.049 | -0.653 | 1.233 | 0.740 |
| How often do you feel isolated from others? | -0.141 | -0.155 | -0.709 | -0.926 | -0.430 | 0.060 |

*Note: S – Standardized discriminant coefficient; F – correlation with discriminant function.*

At the negative side of the first discriminant function (See Figure 1), described dominantly by the lower level of Fear of COVID-19, three centroids of groups lay from less endangered countries (C1); the results of males from C1 countries are either negative or near-zero, values rising from youngest group onwards (Table 3, Figure 1). The centroids of female groups from C1 countries show also that fear is getting higher with age; compared with males, female centroids are shifted to higher values and only the youngest group centroid is positioned at the negative side of function. All the centroids of groups from Italy and Spain (C2) on the first discriminant function are on the positive side of function; they do not show the same regularity as for groups from less endangered countries; the first three age groups of males have lower values then corresponding female groups, but, the eldest males have the highest centroid value on the first function. The emerging female group have the lowest value, whereas the highest value is in young adult female group.

**Table 3.** Centroids distinguished according to country (C1 and C2), gender (M, F) and age (1-emerging adults 18-25 years, 2-young adults 26-39 years, 3 – middle-aged adults 40 – 59 years and 4 - elderly of 60 years and more)

|  | Countries 1 (C1) |  |  | Countries 2 (C2) |  |  |
| --- | --- | --- | --- | --- | --- | --- |
|  | Root 1 | Root 2 | Root 3 | Root 1 | Root 2 | Root 3 |
| M1 | -0.587 | 0.141 | -0.047 | 0.281 | 0.226 | -0.138 |
| M2 | -0.316 | 0.244 | -0.075 | 0.252 | 0.164 | -0.083 |
| M3 | -0.003 | 0.309 | 0.082 | 0.558 | 0.413 | 0.223 |
| M4 | 0.082 | 0.087 | 0.167 | 1.206 | -0.164 | -0.030 |
| F1 | -0.286 | -0.229 | -0.004 | 0.504 | 0.007 | -0.151 |
| F2 | 0.078 | -0.017 | 0.001 | 0.902 | -0.248 | -0.192 |
| F3 | 0.269 | -0.036 | 0.207 | 0.859 | 0.036 | -0.108 |
| F4 | 0.727 | -0.153 | -0.053 | 0.613 | 0.009 | 0.087 |

**Figure 1.**
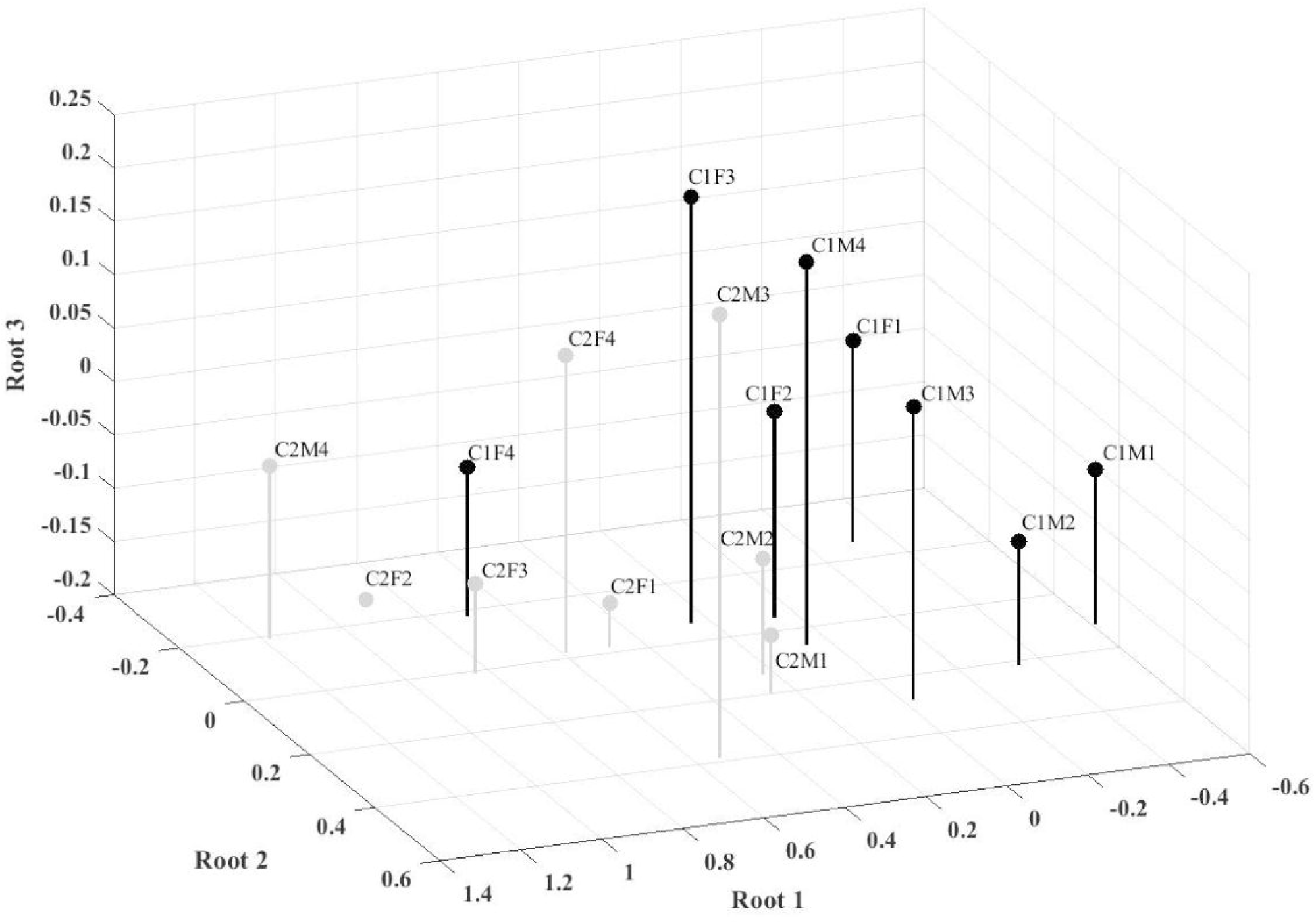
Centroids representation on first, second and third discriminant functions Note: country (Cl and C2), (M, F) and age 1-emerging adults 18-25 years, 2-young adults 26-39 years, 3 - middle-agedlife adults 40 - 59 years and 4 - elderly of 60 years and more)

The centroids of groups from less endangered countries (C1) on the second discriminant function show that females feel more often isolated from others and feel the lack of companionship more than males. Three female groups from Italy and Spain (C2) have near-zero values and only the young adult female group has centroid on the negative side of function; males from the first three age groups have centroids on the positive side of function, and the eldest male group has the centroid on the negative side of the function.

The third discriminant function is defined by the feeling of being left out. In the groups from less endangered countries (C1) only elderly males and middle-aged females have centroids on the positive side of the function, the others have near-zero values. Seven groups from Italy and Spain (C2) have near-zero or centroids on the negative side of the function; only the group of middle-aged males felt more often to be left out.

## Discussion

The present study examined cross-country differences concerning the fear of COVID-19 and loneliness due to varying degrees of outbreak severity. Our results suggest that both males and females in European countries suffering from a strong impact of the COVID-19 pandemic (i.e. Italy and Spain) reported higher fear of COVID-19 and sense of loneliness than countries with a low spread of the virus in April 2020. From the analyses, it resulted that people who had high level of fear of COVID-19 tended to suffer loneliness in a less extent, and who was feeling more isolated, tended also to feel the lack of companionship. However, discriminant analysis showed that this pattern of results can be examined in more detail by considering the different age and gender groups.

As expected, from the analysis of the centroids, in countries with low death rate and mild social restrictions (C1), both the emerging and young adults did not show high level of fear, but felt the lack of companionship, especially for men; whereas middle-aged and older women showed higher level of fear and low lack of companionship. Conversely, people from high death rate countries and harsh restrictions (C2) experienced higher fear of COVID-19, without feeling the lack of companionship. Furthermore, middle-aged men and young adult women felt both the lack of companionship and to be left out. These results are not surprising if we consider that during the time-lag of the survey, Italy and Spain faced strict restrictions and lockdown, which could have impacted on people’s sense of loneliness. Moreover, Italy and Spain faced over 10,000 deaths only in the two weeks of the survey, while Bosnia and Herzegovina, Slovenia, Slovakia, and Serbia had 160 deaths in total. Previous data from the USA also showed that fears appeared to be concentrated in regions with the highest reported COVID-19 cases [24].

Regarding the influence of gender, our results confirmed that women reported greater fear than men both in C1 and C2 countries. This finding is consistent with literature showing that females can be more vulnerable to develop psychosocial distress during the pandemic [5, 7]. Research on the impact of COVID-19 pandemic on men’s and women’s well-being separately is still scarce and there is a need for addressing gender equality in any decision making for the COVID-19 [35]. The findings of the current study suggest that the discriminant functions can be used to identify sub-groups at high risk of distress during the COVID-19 pandemic. The elderly females from countries with low death rate could be considered a group at moderate risk of excessive fear of COVID-19 and lack of companionship. Given the mild restrictions assigned in these countries, this vulnerable group could be supported by improving regular exercising and maintaining a healthy diet pattern, to prevent symptoms of stress during the pandemic.

Of note, in Italy and Spain the older men who are at higher risk for COVID-19 complications represents a class of individuals at risk of high fear of COVID-19 and feelings of social isolation. From a policymaking perspective, more attention should be paid to these vulnerable groups by enhancing on-line health services and support. Moreover, these vulnerable groups should be helped to avoid potential false reports and continuously checking COVID-19 related news to ease their feelings of fear and anxiety.

The COVID-19 outbreak is likely worsening individual’s perception of loneliness by reducing social interactions and contacts. Given that loneliness is a risk to physical and mental health, there was a call for a public health framework for addressing loneliness during COVID-19, especially in older adults[36]. Our results showed that the elderly male group in Italy and Spain felt more often isolated from others and felt the lack of companionship. They also reported higher fear of COVID-19 than other age groups. Thus, they may be identified as a sub-group at high risk of social distress during the COVID-19 pandemic [37]. Physicians could help lonely adults to use social services and community-based organizations, and support them to alleviate loneliness and address essential needs [38]. Moreover, our results showed that females in C1 countries felt more often isolated from others and with a lack of companionship than males. It could be speculated that in countries with mild social restrictions due to the COVID-19 outbreak, women were more engaged with demanding family activities than men and had less opportunities for social interactions, thus feeling lonelier in this difficult time.

### Strengths and limitations of the study

The main strength of the current study is that we examined the impact of the COVID-19 outbreak on fear and sense of loneliness in a large sample of population from different European countries. This study also has several limitations: first, well-educated people are more likely to participate in an on-line survey than less-educated, as confirmed by Smith [41]. Moreover, people from low socioeconomic status might not be equipped with the Internet and IT technology. Finally, using self-report measures could not reflect people’s real opinions and feelings, due to the social desirability [42].

## Conclusions

Overall, our results show that people from European countries with a high number of infections and deaths during the COVID-19 pandemic reported different levels of fear and feelings of loneliness than people from countries with a very low death and infection rates. Moreover, our results highlight that future research on the negative health consequences of the COVID-19 pandemic should examine different age and gender groups separately. The analysis of emotional well-being in groups at risk of mental health issues can help to lessen the long term social and economic costs due to the COVID-19 outbreak, and to integrate behavioral health expertise into public health responses to pandemic [39, 40].

## Data Availability

The datasets generated for this study are available on request to the corresponding author.

### S1 Appendix Descriptive statistics of the results of the survey

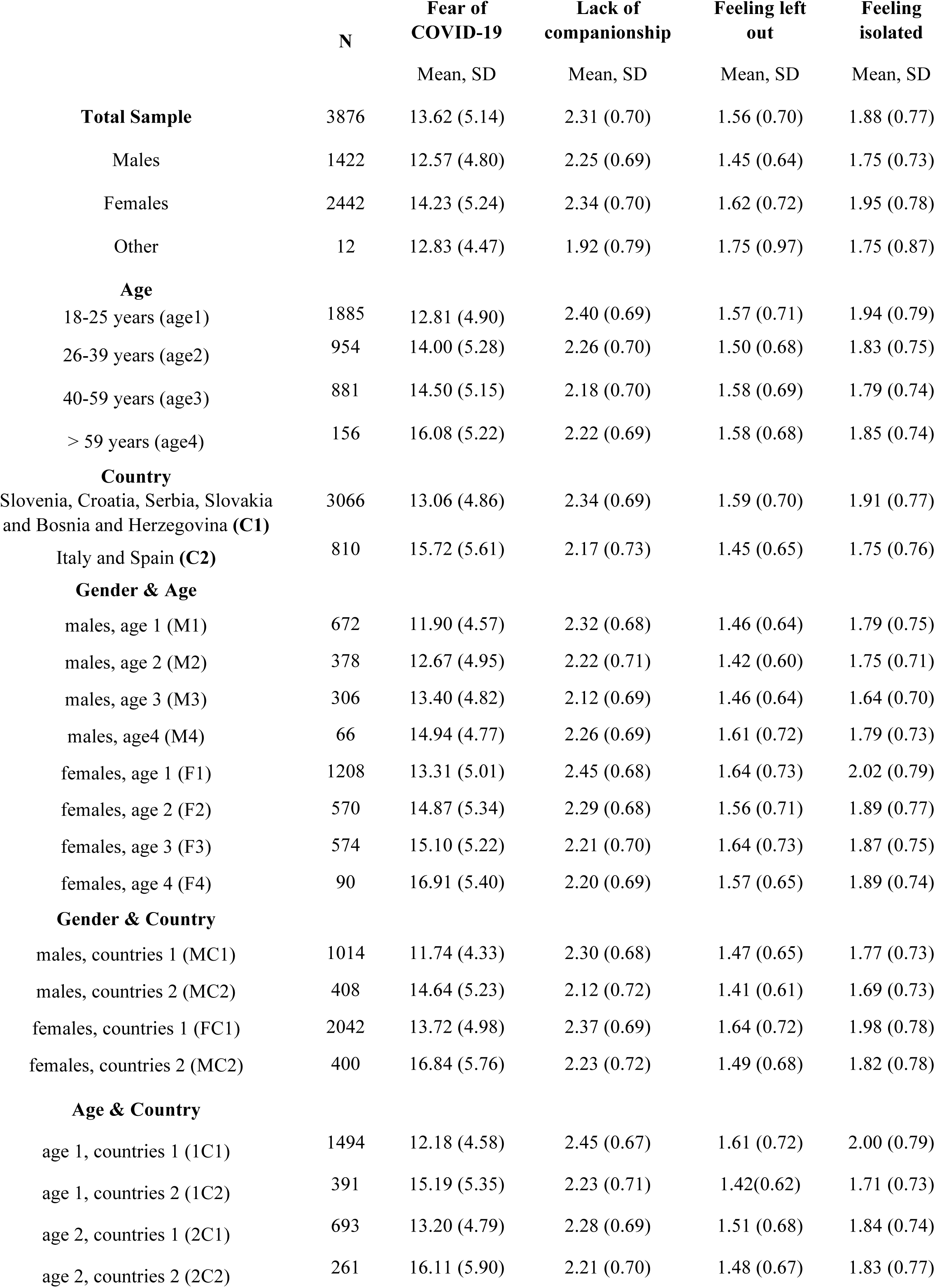
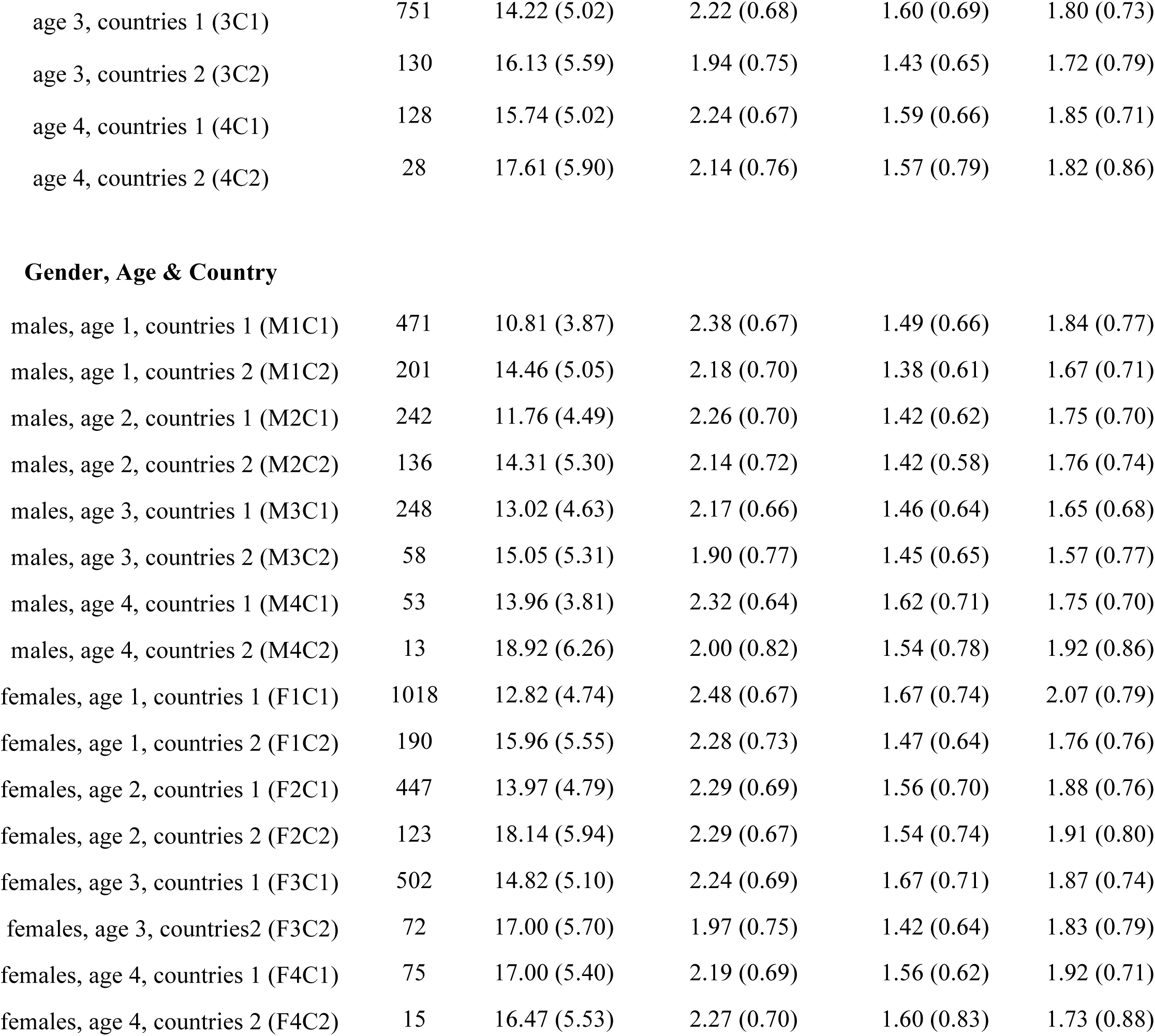

|  | N | Fear of<br>COVID-19 | Lack of<br>companionship | Feeling left<br>out | Feeling<br>isolated |
| --- | --- | --- | --- | --- | --- |
|  |  | Mean, SD | Mean, SD | Mean, SD | Mean, SD |
| **Total Sample** | 3876 | 13.62 (5.14) | 2.31 (0.70) | 1.56 (0.70) | 1.88 (0.77) |
| Males | 1422 | 12.57 (4.80) | 2.25 (0.69) | 1.45 (0.64) | 1.75 (0.73) |
| Females | 2442 | 14.23 (5.24) | 2.34 (0.70) | 1.62 (0.72) | 1.95 (0.78) |
| Other | 12 | 12.83 (4.47) | 1.92 (0.79) | 1.75 (0.97) | 1.75 (0.87) |
| **Age** |  |  |  |  |  |
| 18-25 years (age1) | 1885 | 12.81 (4.90) | 2.40 (0.69) | 1.57 (0.71) | 1.94 (0.79) |
| 26-39 years (age2) | 954 | 14.00 (5.28) | 2.26 (0.70) | 1.50 (0.68) | 1.83 (0.75) |
| 40-59 years (age3) | 881 | 14.50 (5.15) | 2.18 (0.70) | 1.58 (0.69) | 1.79 (0.74) |
| > 59 years (age4) | 156 | 16.08 (5.22) | 2.22 (0.69) | 1.58 (0.68) | 1.85 (0.74) |
| **Country** |  |  |  |  |  |
| Slovenia, Croatia, Serbia, Slovakia<br>and Bosnia and Herzegovina (C1) | 3066 | 13.06 (4.86) | 2.34 (0.69) | 1.59 (0.70) | 1.91 (0.77) |
| Italy and Spain (C2) | 810 | 15.72 (5.61) | 2.17 (0.73) | 1.45 (0.65) | 1.75 (0.76) |
| **Gender & Age** |  |  |  |  |  |
| males, age 1 (M1) | 672 | 11.90 (4.57) | 2.32 (0.68) | 1.46 (0.64) | 1.79 (0.75) |
| males, age 2 (M2) | 378 | 12.67 (4.95) | 2.22 (0.71) | 1.42 (0.60) | 1.75 (0.71) |
| males, age 3 (M3) | 306 | 13.40 (4.82) | 2.12 (0.69) | 1.46 (0.64) | 1.64 (0.70) |
| males, age4 (M4) | 66 | 14.94 (4.77) | 2.26 (0.69) | 1.61 (0.72) | 1.79 (0.73) |
| females, age 1 (F1) | 1208 | 13.31 (5.01) | 2.45 (0.68) | 1.64 (0.73) | 2.02 (0.79) |
| females, age 2 (F2) | 570 | 14.87 (5.34) | 2.29 (0.68) | 1.56 (0.71) | 1.89 (0.77) |
| females, age 3 (F3) | 574 | 15.10 (5.22) | 2.21 (0.70) | 1.64 (0.73) | 1.87 (0.75) |
| females, age 4 (F4) | 90 | 16.91 (5.40) | 2.20 (0.69) | 1.57 (0.65) | 1.89 (0.74) |
| **Gender & Country** |  |  |  |  |  |
| males, countries 1 (MC1) | 1014 | 11.74 (4.33) | 2.30 (0.68) | 1.47 (0.65) | 1.77 (0.73) |
| males, countries 2 (MC2) | 408 | 14.64 (5.23) | 2.12 (0.72) | 1.41 (0.61) | 1.69 (0.73) |
| females, countries 1 (FC1) | 2042 | 13.72 (4.98) | 2.37 (0.69) | 1.64 (0.72) | 1.98 (0.78) |
| females, countries 2 (MC2) | 400 | 16.84 (5.76) | 2.23 (0.72) | 1.49 (0.68) | 1.82 (0.78) |
| **Age & Country** |  |  |  |  |  |
| age 1, countries 1 (1C1) | 1494 | 12.18 (4.58) | 2.45 (0.67) | 1.61 (0.72) | 2.00 (0.79) |
| age 1, countries 2 (1C2) | 391 | 15.19 (5.35) | 2.23 (0.71) | 1.42(0.62) | 1.71 (0.73) |
| age 2, countries 1 (2C1) | 693 | 13.20 (4.79) | 2.28 (0.69) | 1.51 (0.68) | 1.84 (0.74) |
| age 2, countries 2 (2C2) | 261 | 16.11 (5.90) | 2.21 (0.70) | 1.48 (0.67) | 1.83 (0.77) |
| age 3, countries 1 (3C1) | 751 | 14.22 (5.02) | 2.22 (0.68) | 1.60 (0.69) | 1.80 (0.73) |
| age 3, countries 2 (3C2) | 130 | 16.13 (5.59) | 1.94 (0.75) | 1.43 (0.65) | 1.72 (0.79) |
| age 4, countries 1 (4C1) | 128 | 15.74 (5.02) | 2.24 (0.67) | 1.59 (0.66) | 1.85 (0.71) |
| age 4, countries 2 (4C2) | 28 | 17.61 (5.90) | 2.14 (0.76) | 1.57 (0.79) | 1.82 (0.86) |
| Gender, Age & Country |  |  |  |  |  |
| males, age 1, countries 1 (M1C1) | 471 | 10.81 (3.87) | 2.38 (0.67) | 1.49 (0.66) | 1.84 (0.77) |
| males, age 1, countries 2 (M1C2) | 201 | 14.46 (5.05) | 2.18 (0.70) | 1.38 (0.61) | 1.67 (0.71) |
| males, age 2, countries 1 (M2C1) | 242 | 11.76 (4.49) | 2.26 (0.70) | 1.42 (0.62) | 1.75 (0.70) |
| males, age 2, countries 2 (M2C2) | 136 | 14.31 (5.30) | 2.14 (0.72) | 1.42 (0.58) | 1.76 (0.74) |
| males, age 3, countries 1 (M3C1) | 248 | 13.02 (4.63) | 2.17 (0.66) | 1.46 (0.64) | 1.65 (0.68) |
| males, age 3, countries 2 (M3C2) | 58 | 15.05 (5.31) | 1.90 (0.77) | 1.45 (0.65) | 1.57 (0.77) |
| males, age 4, countries 1 (M4C1) | 53 | 13.96 (3.81) | 2.32 (0.64) | 1.62 (0.71) | 1.75 (0.70) |
| males, age 4, countries 2 (M4C2) | 13 | 18.92 (6.26) | 2.00 (0.82) | 1.54 (0.78) | 1.92 (0.86) |
| females, age 1, countries 1 (F1C1) | 1018 | 12.82 (4.74) | 2.48 (0.67) | 1.67 (0.74) | 2.07 (0.79) |
| females, age 1, countries 2 (F1C2) | 190 | 15.96 (5.55) | 2.28 (0.73) | 1.47 (0.64) | 1.76 (0.76) |
| females, age 2, countries 1 (F2C1) | 447 | 13.97 (4.79) | 2.29 (0.69) | 1.56 (0.70) | 1.88 (0.76) |
| females, age 2, countries 2 (F2C2) | 123 | 18.14 (5.94) | 2.29 (0.67) | 1.54 (0.74) | 1.91 (0.80) |
| females, age 3, countries 1 (F3C1) | 502 | 14.82 (5.10) | 2.24 (0.69) | 1.67 (0.71) | 1.87 (0.74) |
| females, age 3, countries2 (F3C2) | 72 | 17.00 (5.70) | 1.97 (0.75) | 1.42 (0.64) | 1.83 (0.79) |
| females, age 4, countries 1 (F4C1) | 75 | 17.00 (5.40) | 2.19 (0.69) | 1.56 (0.62) | 1.92 (0.71) |
| females, age 4, countries 2 (F4C2) | 15 | 16.47 (5.53) | 2.27 (0.70) | 1.60 (0.83) | 1.73 (0.88) |

